# Alcohol use and poor sleep quality: a longitudinal twin study across 36 years

**DOI:** 10.1101/2022.03.11.22272157

**Authors:** Viola Helaakoski, Jaakko Kaprio, Christer Hublin, Hanna M. Ollila, Antti Latvala

## Abstract

Poor sleep is one of the multiple health issues associated with heavy alcohol consumption. While acute effects of alcohol intake on sleep have been widely investigated, the longitudinal associations remain relatively underexplored. The objective of our research was to shed light on cross-sectional and longitudinal associations between alcohol use and insomnia symptoms over time, and to elucidate the role of familial confounding factors in such associations. Using self-report questionnaire data from the Older Finnish Twin Cohort (N=13,851), we examined how alcohol consumption and binge drinking are associated with sleep quality during a period of 36 years. Cross-sectional logistic regression analyses revealed significant associations between poor sleep and alcohol misuse, including heavy and binge drinking, at all four time points (OR range =1.61-3.37, P <0.05), suggesting that higher alcohol intake is associated with poor sleep quality over the years. Longitudinal cross-lagged analyses indicated that moderate, heavy and binge drinking predict poor sleep quality (OR range =1.25-1.76, P <0.05), but not the reverse. Within-pair analyses suggested that the associations between heavy drinking and poor sleep quality were not fully explained by genetic and environmental influences shared by the co-twins. In conclusion, our findings support previous literature in that alcohol use is associated with poor sleep quality, such that alcohol use predicts poor sleep quality later in life, but not vice versa, and that the association is not fully explained by familial factors.

## Introduction

Sleep is a vital, often undervalued, element of mental and physical health. Sleeping problems have become increasingly common among the general population, with insomnia affecting approximately 20% of the general population on an acute, recurring or chronic basis [1-3]. Insomnia is one of the most common reasons for sleeping problems, and is characterised by difficulties with falling asleep, staying asleep or waking up too early, in addition to complaints of nonrestorative sleep. Insomnia generally results in some form of daytime impairment through e.g. fatigue and mood changes. Chronic insomnia is associated with an increased risk of various cardiovascular, autoimmune and psychiatric diseases [4-6]. The causes of insomnia are multifactorial, ranging from genetic influences to different lifestyle factors. A major lifestyle factor associated with insomnia symptoms is alcohol misuse, including heavy alcohol consumption and binge drinking [7]. Because many of the brain systems and neurotransmitters associated with sleep-wake regulation are also influenced by alcohol intake, it is biologically credible that drinking affects sleep [8].

Some individuals ‘self-medicate’ their sleep problems with alcohol [9], even though the negative effects of a single pre-bedtime dose of alcohol on sleep are well known. Previous research suggests that alcohol, through its sedative effect, initially shortens sleep onset latency at all dosages, but disrupts the quality of sleep in the second half of the night. High doses of alcohol appear to significantly suppress rapid eye movement (REM) sleep in the first half of the night, and the total amount of REM sleep is decreased at moderate and high doses. The onset of the first REM sleep stage is considerably delayed at all doses. Alcohol at all dosages also reduces wakefulness in the first half of sleep, but increases it in the second part of the night in accordance with the metabolic elimination of ethanol from the body [7,10].

Previous literature has mainly focused on short-term effects of alcohol on sleep quality, whereas little is known about the long-term effects. Importantly, it is yet to be established whether heavy drinking predicts sleep disruption, or whether it is sleep disruption that increases the risk of alcohol misuse, or whether both effects exist.

Regarding alcohol use disorder (AUD), research consistently indicates a high comorbidity with insomnia, with a large proportion of AUD patients reporting insomnia symptoms either whilst drinking or during recovery [11,12]. It is common for individuals with AUD to develop poor sleep hygiene with an irregular sleep schedule, napping during the day and greater wakefulness at night. On the other hand, poor sleeping habits and insomnia symptoms may in some cases precede AUD, thereby suggesting a potential bidirectional relationship between insomnia and AUD [13]. It should be noted, though, that the development of AUD is typically a result of long-term heavy drinking, and so sleep problems might develop during the subclinical phase of AUD. The chronic effects and directionality between sleep disruption and alcohol use remains less clear in healthy individuals.

The aim of the current research was to expand our understanding of the relationship between alcohol misuse and insomnia symptoms. We investigated how heavy alcohol consumption and binge drinking during adulthood are associated with poor sleep quality across a period of 36 years. Understanding these associations is crucial for developing effective preventive and treatment strategies for both sleep and alcohol disorders.

## Methods

### Data

We used data from the Older Finnish Twin Cohort [14], which includes Finnish monozygotic (MZ) and same-sex dizygotic (DZ) twin pairs born before the year of 1958. The baseline survey took place in 1975, with three follow-up health and lifestyle surveys in 1981, 1990 and 2011. The 1990 survey was completed by twins born in 1930-1957, and the 2011 by twins born in 1945-1957. Only participants born in 1945-1957 were included in our analyses (N = 13,851), so as to control for age differences and exclude those who did not have a chance to take part in all four surveys. Ethical permissions for the Older Twin Cohort surveys were obtained from the Ethics Committee of the Department of Public Health, University of Helsinki and the Ethical Committee of the University Hospital District of Helsinki and Uusimaa.

### Measures

As for sleep variables, we focused on poor sleep quality. In addition, we examined associations with short sleep duration as a sensitivity analysis, as short sleep correlates both with sleep quality and insomnia symptoms. Trends in sleep quality and duration over time in the same cohort have already been extensively investigated [15,16]. The items included in all four surveys, with the distributions for each trait at each time point are reported in Table 1 below. Sleep quality was measured by asking participants whether they tend to sleep: “well”, “fairly well”, “fairly poorly” or “poorly”. For the purposes of our analyses, cases of poor sleep were coded to include those who reported sleeping either “fairly poorly” or “poorly”, whilst those reporting sleeping “well” or “fairly well” were coded as sleeping well. Sleep duration was coded as short sleep (<7 hours), average sleep (7-8 hours) and long sleep (>8 hours). Analyses of short sleep duration were restricted to those reporting short or average sleep, whilst long sleepers were excluded from the analyses.

**Table 1.**
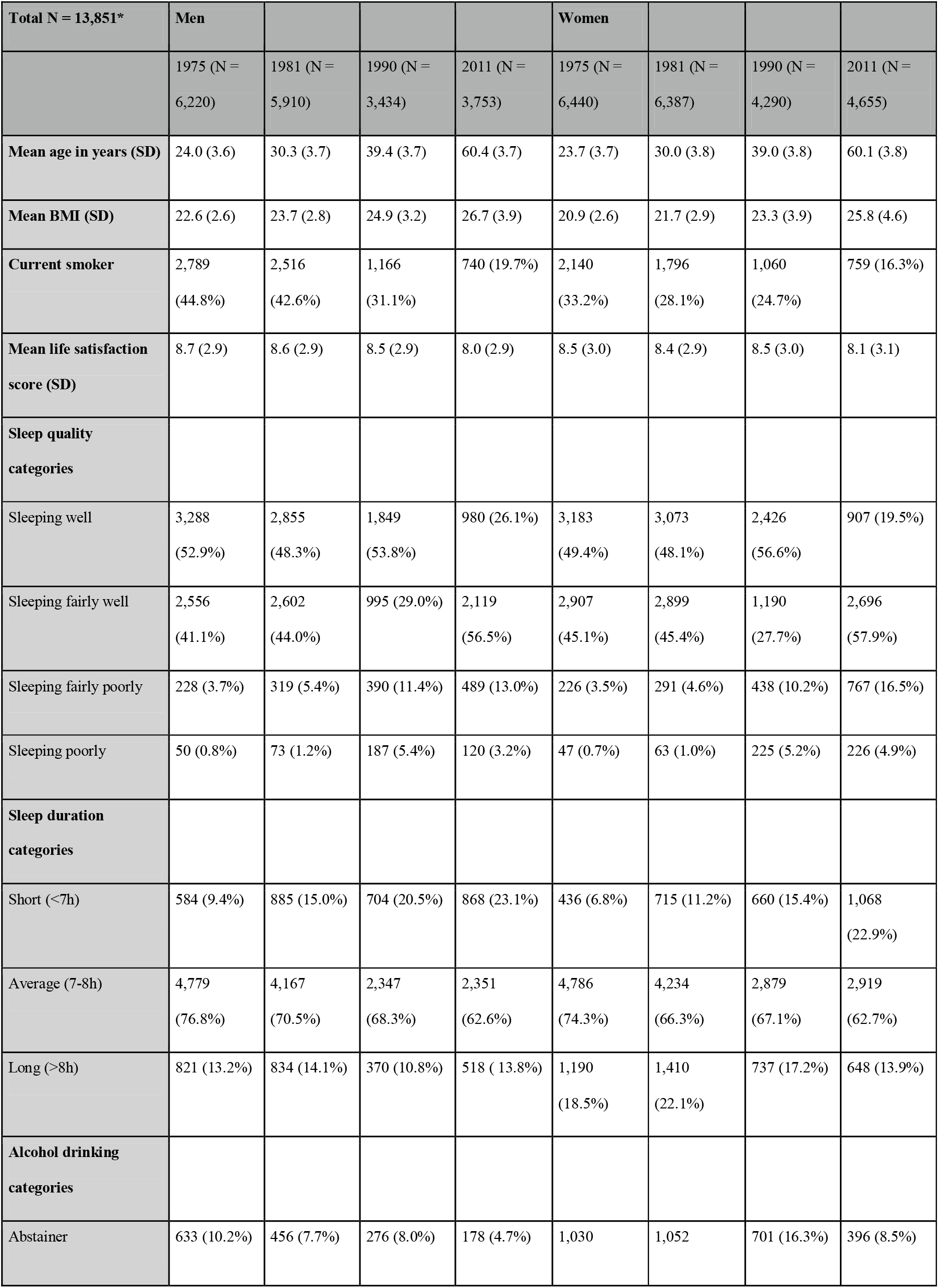
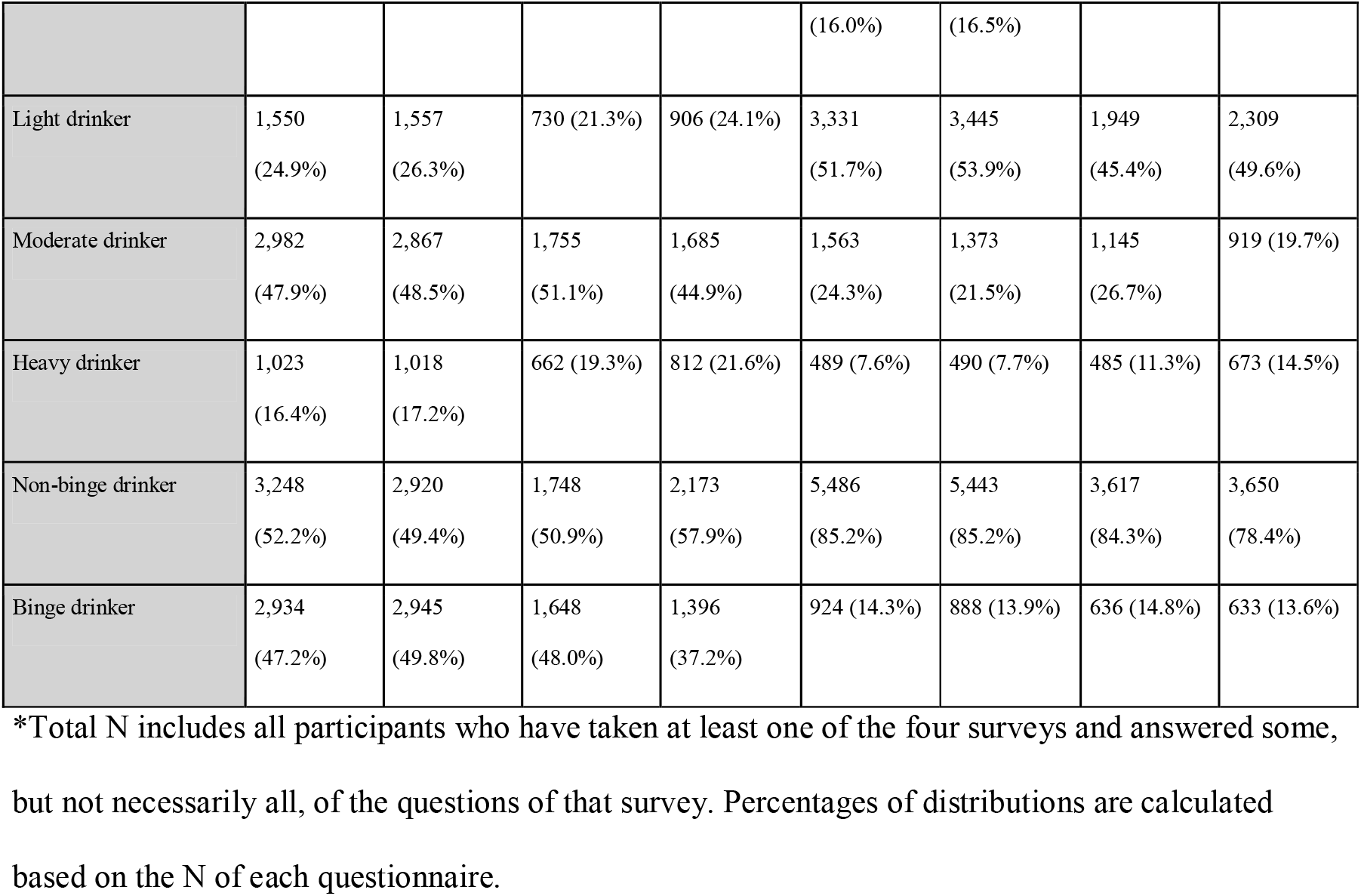
Participant characteristics and distributions of relevant sleep and drinking categories.

Regarding alcohol measures, we investigated the amount of alcohol intake as well as binge drinking. Trends in drinking categories in the Older Finnish Twin Cohort have also been investigated by previous research [17]. Alcohol intake was measured by the monthly amount of alcohol consumed (in grams). Participants were categorised as “heavy drinkers”, “moderate drinkers”, “light drinkers” and “abstainers”. Drinking levels were defined as per the NIAAA guidelines [18]. Heavy drinkers include those who reported drinking more than 7 drinks per week (336 g/month) for women and more than 14 drinks per week (672 g/month) for men. Moderate drinkers include participants drinking more than 3 drinks but less than 7 drinks per week (145 - 336 g/month) for women and less than 14 drinks per week (145 - 672 g/month) for men. Light drinkers refer to those drinking less than 3 drinks per week (1 - 144 g/month). Finally, abstainers refer to participants who reported consuming no alcohol at all at the time of taking each survey. As we were interested in increased alcohol use, we used light drinking as a baseline category. As for binge drinking, the participants were asked whether they drank more than 5 bottles of beer, 1 bottle of wine or 4 drinks (≥18 millilitres of spirits) on the same occasion at least once a month. Those who were defined as abstainers at the time of taking each survey were omitted from the analyses, because the association between sleep and abstinence is likely to reflect different factors compared to the amount of alcohol consumed (e.g., chronic illness, long-term medication or past AUD).

### Covariates

We included body mass index (BMI), smoking and life satisfaction as covariates in the analyses. BMI was based on self-reported measures of weight in kilograms and height in centimetres at each survey. Smoking refers to current smoking status (yes/no) at the time of taking each survey.

Life satisfaction (LS) refers to a 4-item summary variable which comprises questions regarding how interesting, happy, easy and lonely the participant rates their life at present. LS has previously been correlated with e.g. depressive symptoms in the same cohort [19]. Associations between LS and affective, anxiety and other psychiatric disorders have also been established in other cohorts [20,21], and so LS can be used as a proxy to estimate overall mental health in self-reported questionnaires where clinical diagnoses are not available. Previous research also suggests that life satisfaction and heavy alcohol consumption predict each other [22], and so adjusting for LS can be used to adjust for potential confounding. The total score ranged from 4 (high LS) to 20 (low LS). The sum score was calculated allowing one missing item. LS was available at each survey.

### Statistical analysis

After running descriptive statistics, we conducted logistic regression analyses to investigate cross-sectional associations between the sleep and alcohol traits. Using cluster correction, we adjusted the analyses for sampling of twin individuals as twin pairs [23]. As for covariates, all analyses were first adjusted for age and sex (Model 1). We then performed the same analyses by including BMI, smoking and LS as additional covariates (Model 2) to rule out possible confounding by these factors. We also conducted interaction analyses to examine sex differences. After this we used a cross-lagged path model to investigate the predictive associations between sleep and alcohol traits over time. In order to have both the alcohol and the sleep variables as binary in the cross-lagged models, we combined the moderate and heavy drinking categories into one and used light drinking as the reference category, similar to the cross-sectional analysis. Finally, we conducted fixed effects within-pair analyses to examine associations within twin pairs, MZ and DZ both separately and together, so as to rule out familial (i.e., genetic and shared environmental) confounding [24].

As an additional analysis, to illuminate the effects of age vs. cohort/period on the cross-sectional associations between alcohol use and sleep measures, we re-ran the analyses for data from the 1975 and 1981 study waves among participants born in 1930-1942 and participants born in 1915-1927, respectively, so that the age ranges matched with the original cohort (i.e., twins born 1945-1957) in the 1990 and 2011 questionnaires.

All analyses were conducted using Stata17 (StataCorp. 2021. *Stata Statistical Software: Release 17.* College Station, TX: StataCorp LLC).

## Results

### Descriptive statistics

Table 1 shows the distributions of the sleep and alcohol variables as well as covariates across study waves. Sleep quality appeared to decline in both genders with age over the years, whereas men appeared to sleep slightly better compared to women. On the other hand, the majority of participants slept well or fairly well throughout the observation period, as has been reported earlier. Sleep duration decreased in both men and women during the 36-year follow-up, whereas most participants reported sleeping 7-8 hours per night at all time points. As for alcohol consumption, drinking quantities increased and the proportion of abstainers decreased over the years. Heavy drinking increased in both genders, whereas trends in binge drinking were somewhat inconsistent. Overall, women appeared to drink substantially less than men.

### Cross-sectional associations between sleep and drinking traits

Including sex and age as covariates (Model 1), cross-sectional analyses between drinking and sleep traits revealed associations between heavy and binge drinking and poor sleep at all time points, with Odds Ratios (ORs) ranging from 1.61 to 3.37 (Table 2). The strongest association was observed between heavy drinking and poor sleep quality in 1981 (OR =3.37 [95% CI: 2.74, 4.14]) when the average age of twins was 30 years. Heavy and binge drinking were also associated with short sleep at all time points. At earlier time points, moderate drinking, as compared to light drinking, was also associated with both poor and short sleep, with ORs ranging from 1.18 to 1.30.

**Table 2.**
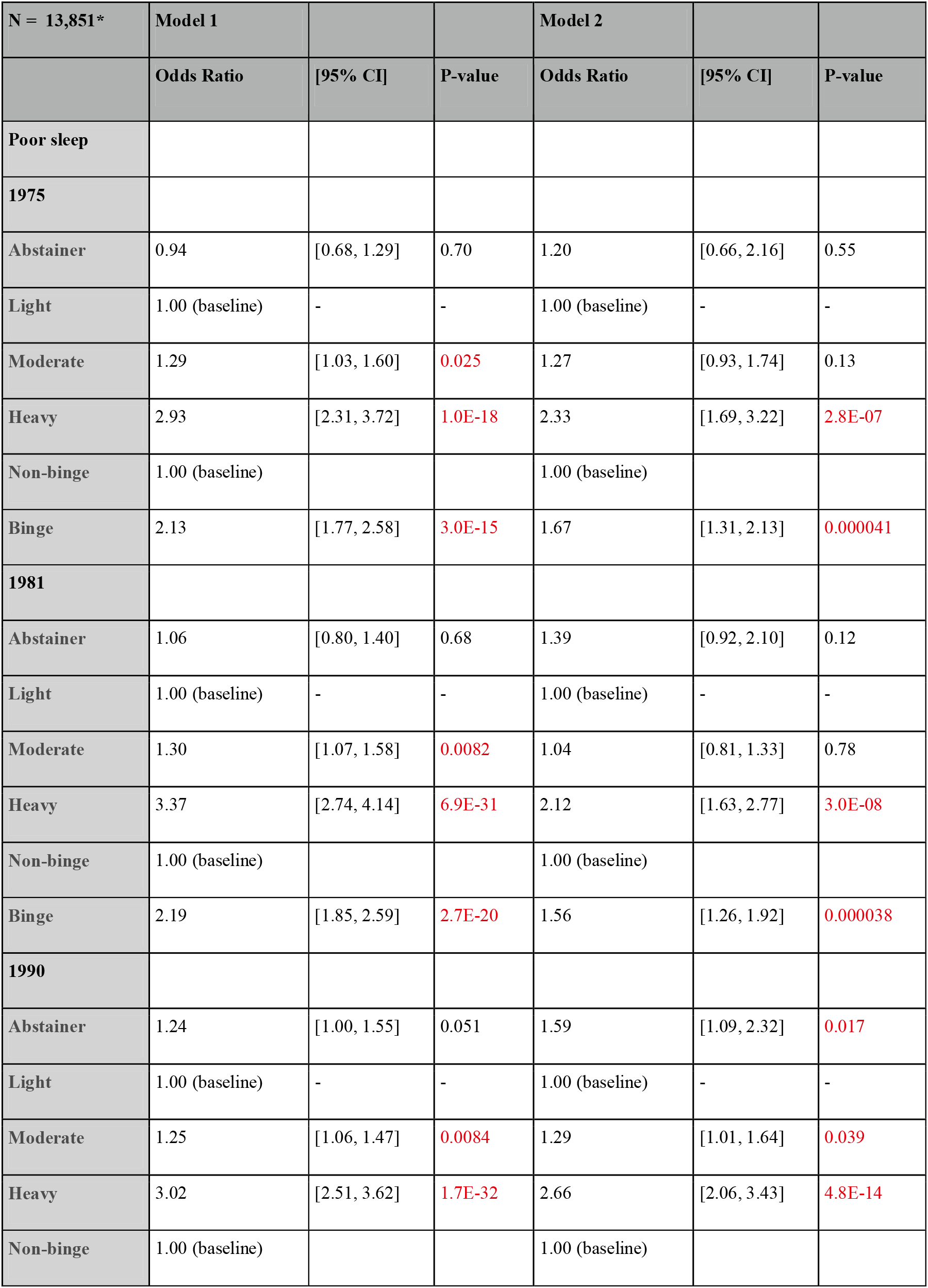
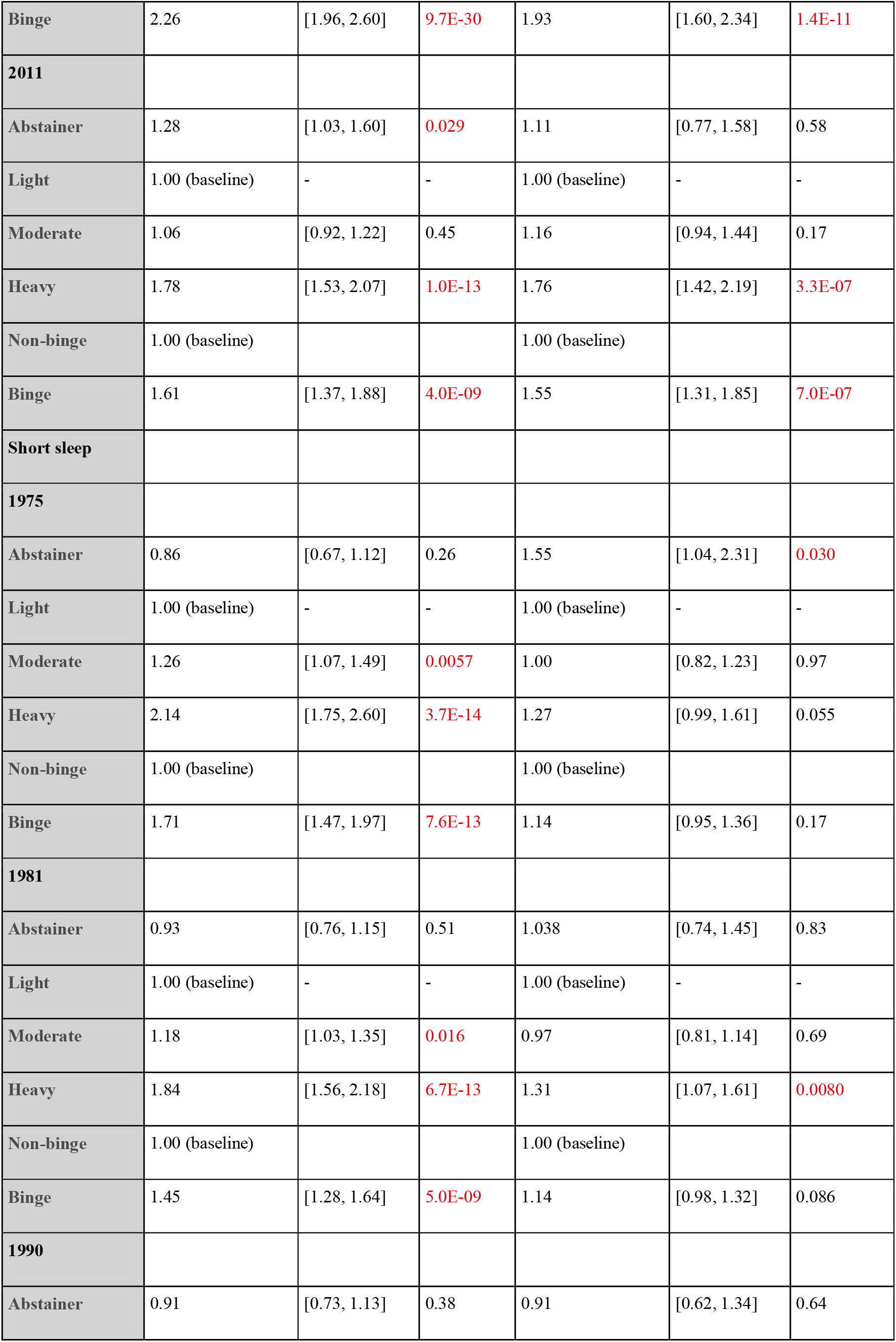
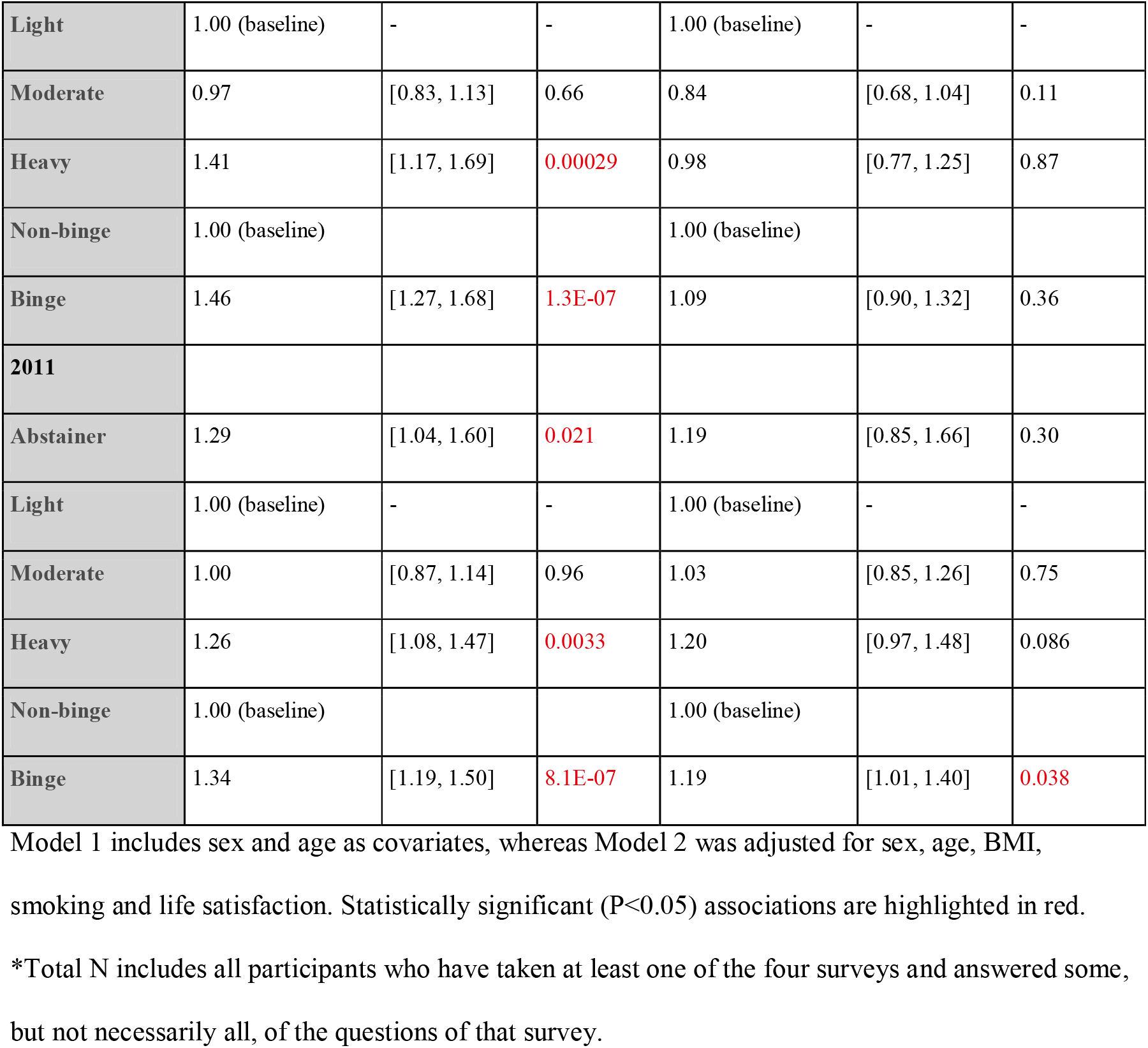
Cross-sectional associations between drinking categories (predictor variables) and sleep traits (outcome variables).

Further adjusting for BMI, smoking and LS (Model 2), associations between heavy/binge drinking and poor sleep quality mostly remained of similar effect sizes as in Model 1, whereas associations with short sleep duration were attenuated (Table 2). Associations between moderate drinking and sleep traits were similarly attenuated. Interaction analyses revealed no statistically significant sex differences in the associations.

In the additional analyses on the effects of age vs. cohort/period on the cross-sectional associations, results from participants born in 1930-1942 in 1975 were statistically less significant with effect sizes being slightly smaller for poor sleep and slightly bigger for short sleep, as compared to results from the main study cohort in 1990. Results from participants born in 1915-1927 in 1981 were non-significant apart from associations with binge drinking where Odds Ratios were slightly smaller for poor sleep and slightly bigger for short sleep, as compared with the original cohort in 2011 (<u>Supplementary Table 1</u>).

### Cross-lagged associations between sleep and drinking traits

Poor sleep at earlier time points predicted poor sleep at later time points across all study waves, and a similar pattern was seen with heavy drinking. Poor sleep was not associated with heavy drinking at subsequent time points, but heavy drinking predicted subsequent poor sleep (Figure 1A). We observed similar, and even slightly stronger associations with binge drinking, so that earlier binge drinking predicted later poor sleep (<u>Supplementary Figure 1A</u>). We then adjusted the models with BMI, smoking and LS assessed at each time point. A very similar pattern from heavy drinking to poor sleep can be seen in both models (Figure 1B). As for binge drinking, the estimates remained consistent after adjusting for other covariates, although not all associations remained statistically significant, suggesting a role of other environmental and lifestyle factors (<u>Supplementary Figure 1B</u>). Results for short sleep were somewhat inconsistent and statistically nonsignificant after adjusting in Model 2 so that no robust conclusions could be drawn.

**Figure 1A.**
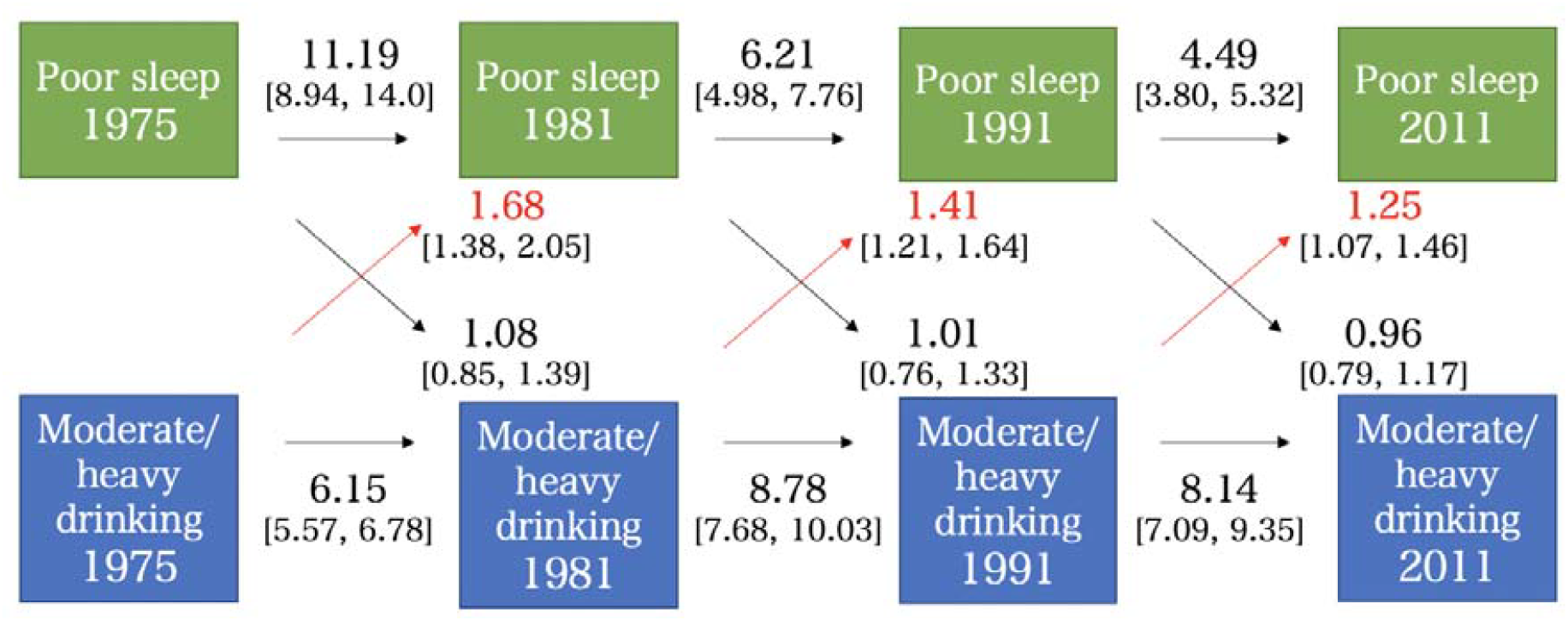
Cross-lagged associations between moderate/heavy drinking and poor sleep quality, adjusting for sex and age (Model 1). The figure shows odds ratios and 95% confidence intervals for each association, with statistically significant (P<0.05) associations between drinking and sleep traits highlighted in red.

**Figure 1B.**
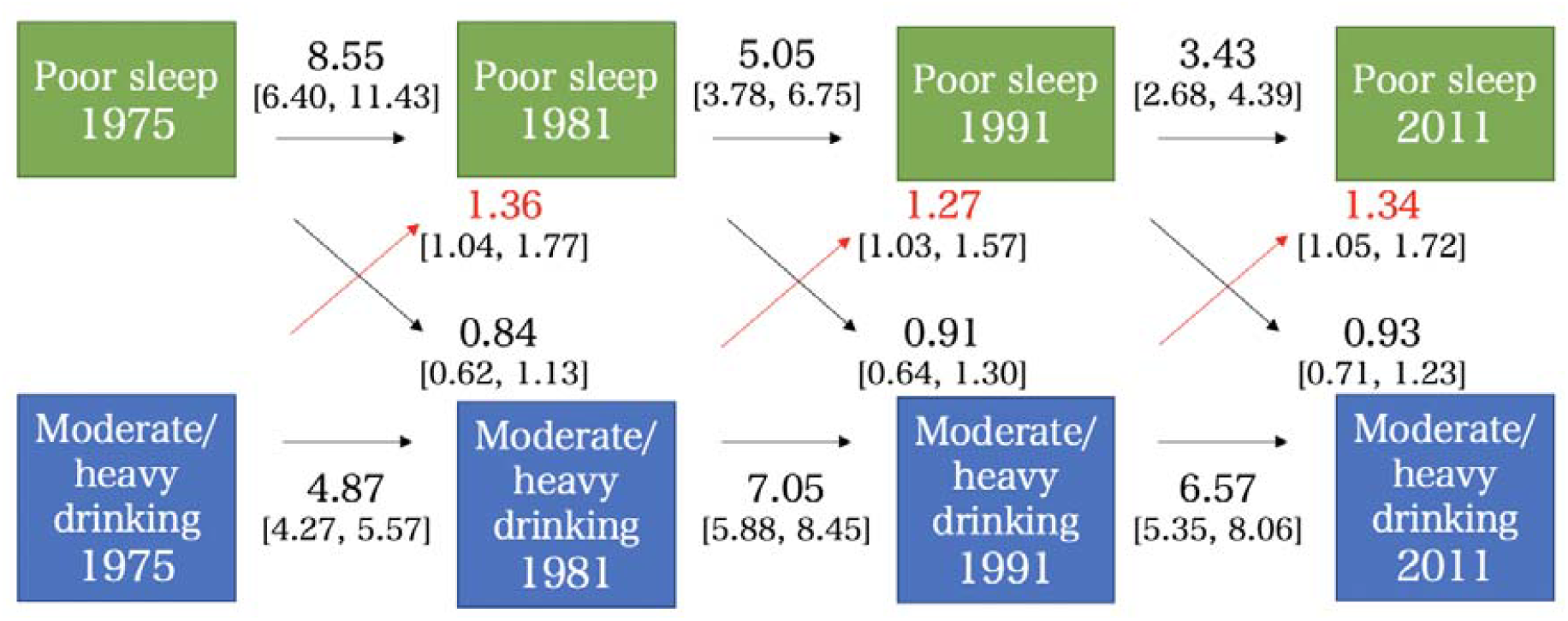
Cross-lagged associations between moderate/heavy drinking and poor sleep quality, adjusting for sex, age, BMI, smoking and life satisfaction (Model 2). The figure shows odds ratios and 95% confidence intervals for each association, with statistically significant (P<0.05) associations between drinking and sleep traits highlighted in red.

### Within-pair models

In within-pair analyses including both MZ and DZ pairs, as shown in Table 3, heavy and binge drinking were associated with an elevated risk of poor sleep quality as compared with light drinking and non-binge drinking (ORs ranging between 1.69 and 2.58), suggesting the associations were not fully explained by familial factors. Overall, associations with short sleep duration were weaker and mostly non-significant. Associations within MZ and DZ twin pairs are given in <u>Supplementary Tables 2A and 2B</u>, respectively. Overall, there were more statistically significant associations within DZ than within MZ pairs, but effect sizes were often of similar magnitude.

**Table 3.** Within-pair associations including both mono- and dizygotic twins.

| **N = 13,851\*** | **Model 1** |  |  | **Model 2** |  |  |
| --- | --- | --- | --- | --- | --- | --- |
|  | **Odds Ratio** | **[95% CI]** | **P-value** | **Odds Ratio** | **[95% CI]** | **P-value** |
| **Poor sleep** |  |  |  |  |  |  |
| **1975** |  |  |  |  |  |  |
| **Abstainer** | 1.12 | [0.66, 1.90] | 0.67 | 1.74 | [0.52, 5.83] | 0.37 |
| **Light** | 1.00 (baseline) |  |  | 1.00 (baseline) |  |  |
| **Moderate** | 0.97 | [0.65, 1.44] | 0.86 | 1.23 | [0.65, 2.32] | 0.52 |
| Heavy | 1.99 | [1.17, 3.38] | 0.011 | 2.12 | [1.00, 4.51] | 0.050 |
| Non-binge | 1.00 (baseline) |  |  | 1.00 (baseline) |  |  |
| Binge | 1.70 | [1.16, 2.47] | 0.0059 | 1.67 | [0.98, 2.85] | 0.061 |
| 1981 |  |  |  |  |  |  |
| Abstainer | 1.39 | [0.81, 2.38] | 0.23 | 2.62 | [0.81, 8.47] | 0.11 |
| Light | 1.00 (baseline) |  |  | 1.00 (baseline) |  |  |
| Moderate | 1.25 | [0.87, 1.78] | 0.23 | 1.72 | [0.97, 3.04] | 0.063 |
| Heavy | 2.04 | [1.31, 3.18] | 0.0016 | 2.58 | [1.24, 5.36] | 0.012 |
| Non-binge | 1.00 (baseline) |  |  | 1.00 (baseline) |  |  |
| Binge | 1.75 | [1.24, 2.47] | 0.0014 | 1.71 | [1.01, 2.91] | 0.046 |
| 1990 |  |  |  |  |  |  |
| Abstainer | 1.28 | [0.83, 1.97] | 0.27 | 0.98 | [0.37, 2.60] | 0.97 |
| Light | 1.00 (baseline) |  |  | 1.00 (baseline) |  |  |
| Moderate | 1.13 | [0.84, 1.52] | 0.43 | 1.22 | [0.73, 2.04] | 0.45 |
| Heavy | 2.65 | [1.78, 3.95] | 1.6E-06 | 2.45 | [1.33, 4.51] | 0.0040 |
| Non-binge | 1.00 (baseline) |  |  | 1.00 (baseline) |  |  |
| Binge | 2.01 | [1.48, 2.72] | 7.0E-06 | 2.05 | [1.27, 3.30] | 0.0033 |
| 2011 |  |  |  |  |  |  |
| Abstainer | 1.35 | [0.87, 2.10] | 0.19 | 1.43 | [0.59, 3.48] | 0.43 |
| Light | 1.00 (baseline) |  |  | 1.00 (baseline) |  |  |
| Moderate | 1.39 | [1.03, 1.87] | 0.032 | 1.31 | [0.75, 2.31] | 0.35 |
| Heavy | 1.74 | [1.23, 2.45] | 0.0016 | 1.80 | [1.01, 3.20] | 0.048 |
| Non-binge | 1.00 (baseline) |  |  | 1.00 (baseline) |  |  |
| Binge | 1.69 | [1.25, 2.30] | 0.00077 | 1.38 | [0.86, 2.24] | 0.19 |
| Short sleep |  |  |  |  |  |  |
| **1975** |  |  |  |  |  |  |
| Abstainer | 1.06 | [0.62, 1.81] | 0.84 | 0.89 | [0.38, 2.11] | 0.80 |
| Light | 1.00 (baseline) |  |  | 1.00 (baseline) |  |  |
| Moderate | 1.28 | [0.93, 1.76] | 0.13 | 1.47 | [0.92, 2.36] | 0.11 |
| Heavy | 2.56 | [1.64, 4.00] | 0.000038 | 3.16 | [1.65, 6.05] | 0.00049 |
| Non-binge | 1.00 (baseline) |  |  | 1.00 (baseline) |  |  |
| Binge | 1.38 | [1.04, 1.84] | 0.026 | 1.00 | [0.69, 1.46] | 0.98 |
| **1981** |  |  |  |  |  |  |
| Abstainer | 0.99 | [0.66, 1.50] | 0.98 | 1.24 | [0.61, 2.52] | 0.55 |
| Light | 1.00 (baseline) |  |  | 1.00 (baseline) |  |  |
| Moderate | 1.16 | [0.89, 1.51] | 0.28 | 0.94 | [0.66, 1.35] | 0.76 |
| Heavy | 1.76 | [1.18, 2.65] | 0.0061 | 1.32 | [0.78, 2.26] | 0.30 |
| Non-binge | 1.00 (baseline) |  |  | 1.00 (baseline) |  |  |
| Binge | 1.25 | [0.95, 1.63] | 0.11 | 1.15 | [0.80, 1.66] | 0.46 |
| **1990** |  |  |  |  |  |  |
| Abstainer | 0.74 | [0.47, 1.18] | 0.21 | 0.36 | [0.091, 1.39] | 0.14 |
| Light | 1.00 (baseline) |  |  | 1.00 (baseline) |  |  |
| Moderate | 1.04 | [0.78, 1.40] | 0.78 | 0.89 | [0.55, 1.43] | 0.62 |
| Heavy | 1.42 | [0.92, 2.19] | 0.11 | 1.11 | [0.59, 2.09] | 0.74 |
| Non-binge | 1.00 (baseline) |  |  | 1.00 (baseline) |  |  |
| Binge | 1.37 | [0.99, 1.90] | 0.055 | 1.54 | [0.98, 2.41] | 0.059 |
| **2011** |  |  |  |  |  |  |
| Abstainer | 1.38 | [0.86, 2.23] | 0.19 | 0.94 | [0.35, 2.51] | 0.90 |
| Light | 1.00 (baseline) |  |  | 1.00 (baseline) |  |  |
| Moderate | 1.17 | [0.89, 1.54] | 0.25 | 1.12 | [0.69, 1.82] | 0.64 |
| **Heavy** | 1.36 | [0.95, 1.93] | 0.091 | 1.14 | [0.64, 2.04] | 0.66 |
| **Non-binge** | 1.00 (baseline) |  |  | 1.00 (baseline) |  |  |
| **Binge** | 1.15 | [0.90, 1.47] | 0.27 | 1.14 | [0.71, 1.84] | 0.58 |

Model 1 includes sex and age as covariates, whereas Model 2 was adjusted for sex, age, BMI,
smoking and life satisfaction. Statistically significant ( $P < 0.05$ ) associations are highlighted in red.
\*Total N includes all participants who have taken at least one of the four surveys and answered some, but not necessarily all, of the questions of that survey.

## Discussion

The findings of the current study confirm that heavy and binge drinking are associated with poor sleep quality. Leveraging repeated measurements across 36 years, cross-sectional analyses reveal that heavy and binge drinking are associated with poor sleep at all time points through adulthood. Our findings not only establish a robust association between high volumes of drinking and poor sleep, but also suggest that moderate and heavy alcohol consumption precede poor sleep in a longitudinal setting, whereas we found little evidence for the reverse associations.

Our results support the numerous earlier findings suggesting that drinking and sleep are connected (reviewed in e.g. [25,26]). The immediate effects of alcohol on sleep are well established, suggesting that consuming even relatively small amounts of alcohol before bedtime increases sleep fragmentation and impairs REM sleep [27,28]. The long-term effects of alcohol on sleep and vice versa require further research. Earlier studies suggest that the relationship of alcohol misuse with sleep is bidirectional. A growing body of literature comprises studies suggesting that insomnia symptoms are associated with subsequent drinking [29,30], whereas other studies have found alcohol use and dependence to precede insomnia [12,31]. Results of the current study are consistent with possible effects of binge and heavy drinking on subsequent poor sleep quality, but not vice versa. Hence, contrary to previous studies, we find that the relationship between drinking and sleep is not bidirectional in nature, although our results cannot establish causality.

Previous literature encompasses few similar longitudinal studies with which to compare our results. There are some population based studies that have investigated longitudinal associations between sleep and alcohol use from adolescence [32] to emerging adulthood [33] and later adulthood [34], but most studies in the field have focused on investigating individuals diagnosed with AUD. We were interested in shedding light on the relationship between sleep and alcohol drinking in the general population in adulthood, so as to better understand the underlying factors influencing the development of both insomnia and AUD.

Using data from twin pairs, we were able to investigate the role of shared familial factors underlying the associations between alcohol use and sleep. Importantly, within-pair analyses suggested that the cross-sectional associations between heavy drinking and poor sleep quality were not fully explained by genetic and environmental influences shared by the co-twins. These findings do not establish a causal effect of alcohol use on sleep quality, but they are a first step in this direction as they enable us to rule out the possibility of a spurious association due to familial background. These findings are novel, as we are not aware of earlier twin studies on the associations between alcohol use and sleep quality. However, earlier research has suggested shared genetic influences on a related sleep trait, namely diurnal preference and problematic alcohol use, including increased quantity and binge drinking [35].

Heavy alcohol consumption and binge drinking are obvious risk factors and potential symptoms of AUD. Our findings suggest that both of these drinking traits are cross-sectionally connected to poor sleep and longitudinally predictive of poor sleep. Even though engaging in risky drinking even over a long period of time does not necessarily equal AUD, it should be noted that the majority of AUD cases remain undiagnosed and thereby untreated [36]. Although we are unable to speculate on the reasons behind drinking in the current study, previous literature suggests that many cases of alcohol consumption can be explained by the self-medication framework [9,37]. When alcohol is used to ‘treat’ personal problems, including sleep problems, symptoms of AUD might develop whilst being left unnoticed. It is well known that chronic misuse of alcohol contributes to severe health consequences. What is often ignored is how drinking contributes to chronic disease by increasing the risk of developing insomnia, other sleep disorders or comorbidities. Accordingly, the health outcomes associated with or mediated by alcohol drinking might be greater than expected by the general population.

All earlier evidence taken together points towards a multifactorial connection between sleep and alcohol including the different components of sleep. Whereas increased drinking appears to be predictive of poor sleep according to our findings, other components of sleep may be differently associated with other patterns and levels of alcohol use. Indeed, in the context of sleep, it is worth noting that different neurotransmitter systems control different aspects of sleep so that timing (chronotype), duration (homeostatic need for sleep) and quality of sleep have partially independent mechanisms [38]. These systems might underlie the reasons why we were unable to see a consistent pattern from increased drinking to short sleep duration similar to that from increased drinking to poor sleep quality.

Our study is among the very few longitudinal studies using a cross-lagged model to investigate associations between insomnia symptoms and alcohol consumption. The model enables us to see how associations develop during the long follow-up period, whilst controlling for associations within different time points and stability across time. It is also worth highlighting the uniqueness of the Older Finnish Twin Cohort data. What makes it scientifically invaluable is not only the fact that it consists of twin participants, but also its long-term follow-up period of 36 years and the comprehensiveness of the questionnaires. The Finnish Twin Cohort is representative of the population with e.g. mortality and cancer incidence being the same as in the general adult population [39,40].

Our findings should be interpreted in the light of the following limitations. Firstly, self-reported data always entails the risk of reporting bias. Secondly, despite the longitudinal setting, this study on its own cannot establish causation and should be interpreted in the light of earlier and accumulating literature.

## Conclusion

It has been well established that not getting enough good quality sleep increases the risk of disease. Our research implies that long-term alcohol drinking in adulthood predicts decreased sleep quality later in life, and that familial factors do not fully account for this association. We wish to highlight that consuming moderate and large amounts of alcohol might adversely influence the quality of sleep over time, thereby increasing the risk of developing chronic sleep problems and affecting overall health.

## Data Availability

Persons interested in accessing the data should contact the corresponding author.

## Acknowledgments

The authors would like to thank all participants of the Older Finnish Twin Cohort for their time and interest in advancing scientific research.

## Data Availability

Persons interested in accessing the data should contact the corresponding author.

