## Supplementary Material for "Alcohol use and poor sleep quality: a longitudinal twin study across 36 years"

**Supplementary Figure 1A**. Cross-lagged associations between binge drinking and poor sleep quality, adjusting for sex and age (Model 1).

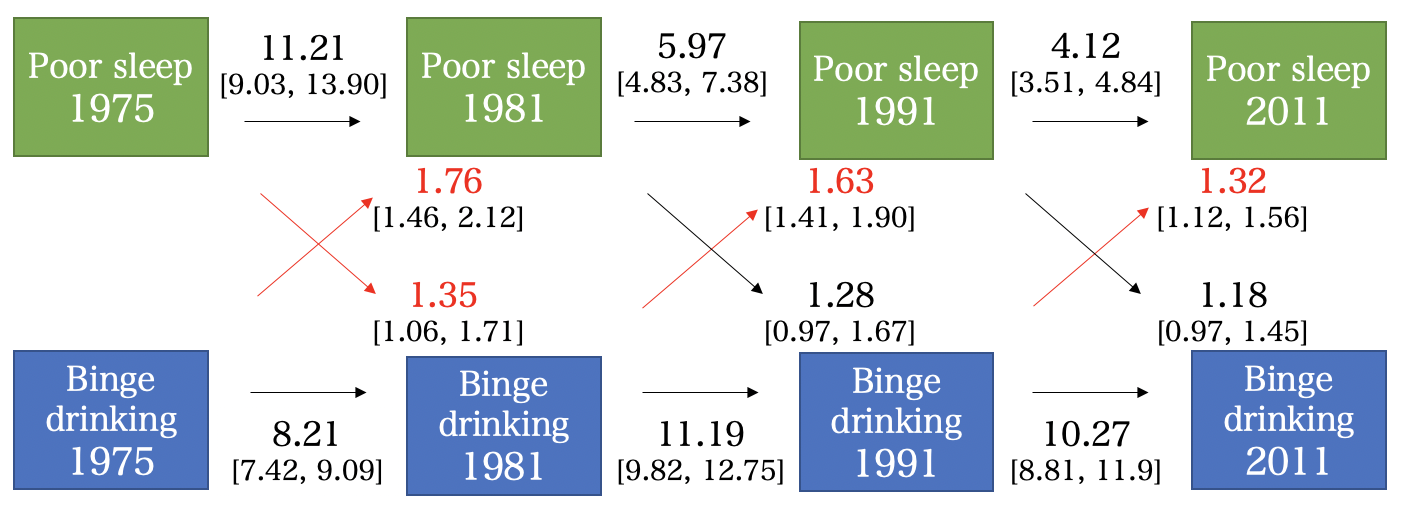

The figure shows odds ratios and 95% confidence intervals in square brackets for each association, with statistically significant (P<0.05) associations between drinking and sleep traits highlighted in red.

**Supplementary Figure 1B**. Cross-lagged associations between binge drinking and poor sleep quality, adjusting for sex, age, BMI, smoking and life satisfaction (Model 2).

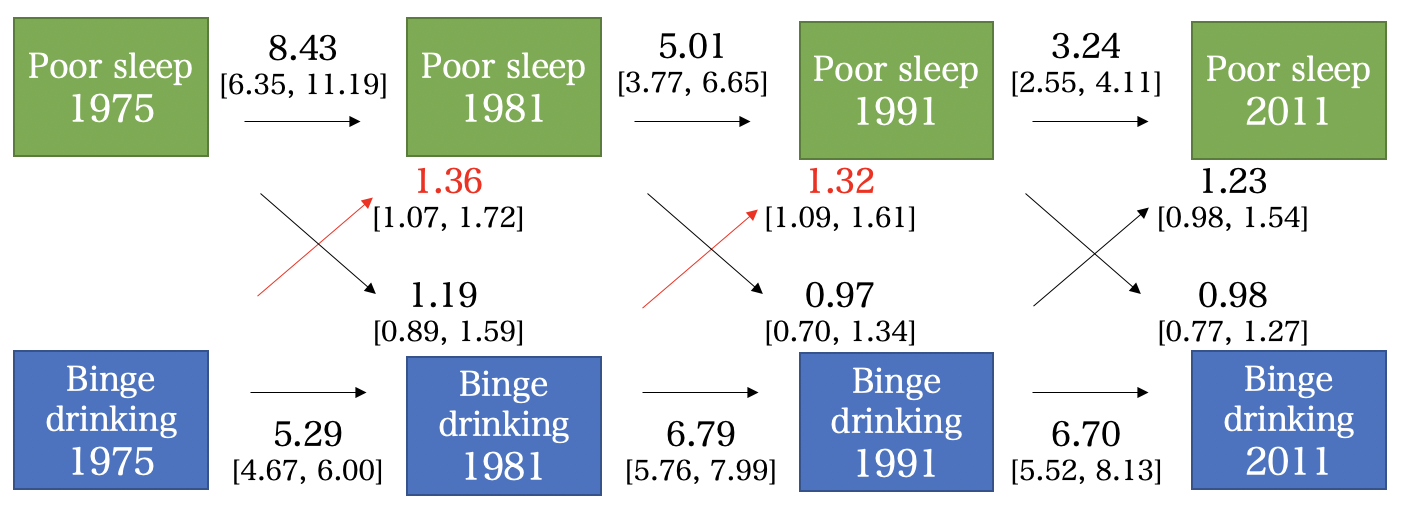

The figure shows odds ratios and 95% confidence intervals for each association, with statistically significant (P<0.05) associations between drinking and sleep traits highlighted in red.

**Supplementary Table 1.** Cross-sectional associations between drinking (predictor variables) and sleep categories (outcome variables).

| **N =  6,191 in 1975*** | **Model 1** |  |  | **Model 2** |  |  |
| --- | --- | --- | --- | --- | --- | --- |
| **N =  3,058 in 1981**** | **Odds Ratio** | **[95% CI]** | **P-value** | **Odds Ratio** | **[95% CI]** | **P-value** |
| **Poor sleep** |  |  |  |  |  |  |
| **1975** |  |  |  |  |  |  |
| **Abstainer** | 1.20 | [0.91, 1.59] | 0.20 | 1.39 | [0.74, 2.59] | 0.30 |
| **Light** | 1.00 (baseline) |  |  | 1.00 (baseline) |  |  |
| **Moderate** | 1.20 | [0.94, 1.53] | 0.15 | 1.24 | [0.86, 1.79] | 0.24 |
| **Heavy** | 2.67 | [2.03, 3.50] | 1.6E-12 | 2.26 | [1.56, 3.28] | 0.000015 |
| **Non-binge** | 1.00 (baseline) |  |  | 1.00 (baseline) |  |  |
| **Binge** | 1.99 | [1.61, 2.47] | 3.2E-10 | 1.52 | [1.15, 2.00] | 0.0033 |
| **1981** |  |  |  |  |  |  |
| **Abstainer** | 1.02 | [0.81, 1.28] | 0.86 | 0.91 | [0.58, 1.42] | 0.67 |
| **Light** | 1.00 (baseline) |  |  | 1.00 (baseline) |  |  |
| **Moderate** | 0.85 | [0.63, 1.14] | 0.27 | 0.71 | [0.49, 1.02] | 0.067 |
| **Heavy** | 1.38 | [0.93, 2.05] | 0.12 | 1.15 | [0.70, 1.89] | 0.58 |
| **Non-binge** | 1.00 (baseline) |  |  | 1.00 (baseline) |  |  |
| **Binge** | 1.50 | [1.08, 2.08] | 0.016 | 1.28 | [0.86, 1.90] | 0.23 |
| **Short sleep** |  |  |  |  |  |  |
| **1975** |  |  |  |  |  |  |
| **Abstainer** | 1.06 | [0.81, 1.39] | 0.68 | 1.45 | [0.87, 2.43] | 0.16 |
| **Light** | 1.00 (baseline) |  |  | 1.00 (baseline) |  |  |
| **Moderate** | 1.31 | [1.05, 1.64] | 0.016 | 1.11 | [0.82, 1.48] | 0.50 |
| **Heavy** | 1.71 | [1.31, 2.24] | 0.000080 | 1.15 | [0.82, 1.60] | 0.41 |
| **Non-binge** | 1.00 (baseline) |  |  | 1.00 (baseline) |  |  |
| **Binge** | 1.60 | [1.30, 1.97] | 0.000011 | 1.24 | [0.97, 1.58] | 0.090 |
| **1981** |  |  |  |  |  |  |
| **Abstainer** | 0.99 | [0.78, 1.26] | 0.96 | 1.14 | [0.74, 1.73] | 0.56 |
| **Light** | 1.00 (baseline) |  |  | 1.00 (baseline) |  |  |
| **Moderate** | 0.96 | [0.74, 1.26] | 0.77 | 0.83 | [0.59, 1.17] | 0.29 |
| **Heavy** | 1.32 | [0.87, 1.99] | 0.19 | 1.14 | [0.69, 1.87] | 0.61 |
| **Non-binge** | 1.00 (baseline) |  |  | 1.00 (baseline) |  |  |
| **Binge** | 1.39 | [1.01, 1.92] | 0.043 | 1.17 | [0.82, 1.68] | 0.39 |

Model 1 includes sex and age as covariates, whereas Model 2 was adjusted for sex, age, BMI, smoking and life satisfaction. Statistically significant (P<0.05) associations are highlighted in red.

*1975 associations were analysed using participants born between 1930-1942.

**1981 associations were analysed using participants born between 1915-1927.

**Supplementary Table 2A.** Within-pair associations in monozygotic twins.

| **N =  3,957*** | **Model 1** |  |  | **Model 2** |  |  |
| --- | --- | --- | --- | --- | --- | --- |
|  | **Odds Ratio** | **[95% CI]** | **P-value** | **Odds Ratio** | **[95% CI]** | **P-value** |
| **Poor sleep** |  |  |  |  |  |  |
| **1975** |  |  |  |  |  |  |
| **Abstainer** | 0.91 | [0.30, 2.70] | 0.86 | 0.94 | [0.14, 6.43] | 0.95 |
| **Light** | 1.00 (baseline) |  |  | 1.00 (baseline) |  |  |
| **Moderate** | 2.63 | [0.99, 7.00] | 0.053 | 2.46 | [0.62, 9.80] | 0.20 |
| **Heavy** | 3.16 | [1.07, 9.33] | 0.037 | 2.93 | [0.78, 10.97] | 0.11 |
| **Non-binge** | 1.00 (baseline) |  |  | 1.00 (baseline) |  |  |
| **Binge** | 1.31 | [0.64, 2.69] | 0.47 | 0.86 | [0.34, 2.15] | 0.74 |
| **1981** |  |  |  |  |  |  |
| **Abstainer** | 0.89 | [0.32, 2.42] | 0.81 | 12.41 | [0.64, 239.52] | 0.095 |
| **Light** | 1.00 (baseline) |  |  | 1.00 (baseline) |  |  |
| **Moderate** | 2.23 | [0.99, 5.05] | 0.054 | 6.44 | [1.58, 26.20] | 0.0093 |
| **Heavy** | 2.63 | [1.00, 6.91] | 0.050 | 7.33 | [1.45, 37.09] | 0.016 |
| **Non-binge** | 1.00 (baseline) |  |  | 1.00 (baseline) |  |  |
| **Binge** | 1.71 | [0.89, 3.25] | 0.11 | 1.85 | [0.76, 4.52] | 1.77 |
| **1990** |  |  |  |  |  |  |
| **Abstainer** | 0.87 | [0.37, 2.04] | 0.76 | 0.66 | [0.089, 4.95] | 0.69 |
| **Light** | 1.00 (baseline) |  |  | 1.00 (baseline) |  |  |
| **Moderate** | 1.35 | [0.75, 2.41] | 0.32 | 0.89 | [0.34, 2.37] | 0.82 |
| **Heavy** | 2.67 | [1.22, 5.84] | 0.014 | 1.96 | [0.58, 6.58] | 0.28 |
| **Non-binge** | 1.00 (baseline) |  |  | 1.00 (baseline) |  |  |
| **Binge** | 2.00 | [1.08, 3.72] | 0.028 | 1.80 | [0.73, 4.45] | 0.20 |
| **2011** |  |  |  |  |  |  |
| **Abstainer** | 0.54 | [0.24, 1.23] | 0.14 | 0.50 | [0.10, 2.47] | 0.39 |
| **Light** | 1.00 (baseline) |  |  | 1.00 (baseline) |  |  |
| **Moderate** | 0.90 | [0.50, 1.63] | 0.74 | 0.81 | [0.31, 2.11] | 0.66 |
| **Heavy** | 0.80 | [0.39, 1.60] | 0.52 | 0.87 | [0.25, 3.01] | 0.83 |
| **Non-binge** | 1.00 (baseline) |  |  | 1.00 (baseline) |  |  |
| **Binge** | 1.87 | [0.92, 3.79] | 0.083 | 0.92 | [0.31, 2.71] | 0.88 |
| **Short sleep** |  |  |  |  |  |  |
| **1975** |  |  |  |  |  |  |
| **Abstainer** | 0.40 | [0.11, 1.50] | 0.17 | 0.53 | [0.096, 2.88] | 0.46 |
| **Light** | 1.00 (baseline) |  |  | 1.00 (baseline) |  |  |
| **Moderate** | 1.92 | [1.00, 3.66] | 0.050 | 3.20 | [1.11, 9.20] | 0.031 |
| **Heavy** | 3.69 | [1.42, 9.55] | 0.0073 | 8.25 | [1.81, 37.66] | 0.0062 |
| **Non-binge** | 1.00 (baseline) |  |  | 1.00 (baseline) |  |  |
| **Binge** | 1.45 | [0.82, 2.56] | 0.20 | 1.18 | [0.57, 2.45] | 0.65 |
| **1981** |  |  |  |  |  |  |
| **Abstainer** | 0.92 | [0.41, 2.07] | 0.85 | 0.79 | [0.18, 3.47] | 0.76 |
| **Light** | 1.00 (baseline) |  |  | 1.00 (baseline) |  |  |
| **Moderate** | 0.89 | [0.52, 1.54] | 0.68 | 0.56 | [0.25, 1.23] | 0.15 |
| **Heavy** | 1.14 | [0.54, 2.44] | 0.73 | 0.64 | [0.23, 1.83] | 0.41 |
| **Non-binge** | 1.00 (baseline) |  |  | 1.00 (baseline) |  |  |
| **Binge** | 0.97 | [0.54, 1.74] | 0.92 | 0.67 | [0.30, 1.51] | 0.34 |
| **1990** |  |  |  |  |  |  |
| **Abstainer** | 0.66 | [0.31, 1.41] | 0.28 | N/A |  |  |
| **Light** | 1.00 (baseline) |  |  | 1.00 (baseline) |  |  |
| **Moderate** | 1.19 | [0.70, 2.00] | 0.52 | 1.66 | [0.71, 3.90] | 0.24 |
| **Heavy** | 1.50 | [0.64, 3.50] | 0.35 | 2.94 | [0.78, 11.05] | 0.11 |
| **Non-binge** | 1.00 (baseline) |  |  | 1.00 (baseline) |  |  |
| **Binge** | 1.84 | [0.97, 3.47] | 0.060 | 2.66 | [1.04, 6.78] | 0.041 |
| **2011** |  |  |  |  |  |  |
| **Abstainer** | 0.93 | [0.38, 2.28] | 0.88 | 0.27 | [0.024, 2.91] | 0.28 |
| **Light** | 1.00 (baseline) |  |  | 1.00 (baseline) |  |  |
| **Moderate** | 0.82 | [0.49, 1.39] | 0.47 | 0.84 | [0.32, 2.23] | 0.73 |
| **Heavy** | 0.81 | [0.41, 1.59] | 0.54 | 0.65 | [0.19, 2.16] | 0.48 |
| **Non-binge** | 1.00 (baseline) |  |  | 1.00 (baseline) |  |  |
| **Binge** | 0.76 | [0.42, 1.38] | 0.37 | 0.83 | [0.32, 2.17] | 0.71 |

Model 1 includes sex and age as covariates, whereas Model 2 was adjusted for sex, age, BMI, smoking and life satisfaction. Statistically significant (P<0.05) associations are highlighted in red.

*Total N includes all monozygotic twin participants who have taken at least one of the four surveys and answered some, but not necessarily all, of the questions of that survey.

**Supplementary Table 2B.** Within-pair associations in dizygotic twins.

| **N =  8,549*** | **Model 1** |  |  | **Model 2** |  |  |
| --- | --- | --- | --- | --- | --- | --- |
|  | **Odds Ratio** | **[95% CI]** | **P-value** | **Odds Ratio** | **[95% CI]** | **P-value** |
| **Poor sleep** |  |  |  |  |  |  |
| **1975** |  |  |  |  |  |  |
| **Abstainer** | 1.19 | [0.64, 2.18] | 0.59 | 3.12 | [0.53, 18.26] | 0.21 |
| **Light** | 1.00 (baseline) |  |  | 1.00 (baseline) |  |  |
| **Moderate** | 0.77 | [0.49, 1.21] | 0.25 | 1.09 | [0.52, 2.30] | 0.82 |
| **Heavy** | 1.91 | [1.01, 3.61] | 0.048 | 2.18 | [0.82, 5.78] | 0.12 |
| **Non-binge** | 1.00 (baseline) |  |  | 1.00 (baseline) |  |  |
| **Binge** | 1.87 | [1.20, 2.91] | 0.0065 | 2.63 | [1.28, 5.41] | 0.0086 |
| **1981** |  |  |  |  |  |  |
| **Abstainer** | 1.68 | [0.87, 3.25] | 0.12 | 2.03 | [0.49, 8.35] | 0.33 |
| **Light** | 1.00 (baseline) |  |  | 1.00 (baseline) |  |  |
| **Moderate** | 1.06 | [0.70, 1.60] | 0.78 | 1.15 | [0.58, 2.28] | 0.70 |
| **Heavy** | 2.03 | [1.22, 3.39] | 0.0065 | 1.94 | [0.81, 4.66] | 0.14 |
| **Non-binge** | 1.00 (baseline) |  |  | 1.00 (baseline) |  |  |
| **Binge** | 1.77 | [1.18, 2.65] | 0.0057 | 1.62 | [0.81, 3.21] | 0.17 |
| **1990** |  |  |  |  |  |  |
| **Abstainer** | 1.43 | [0.86, 2.38] | 0.17 | 1.25 | [0.39, 3.97] | 0.71 |
| **Light** | 1.00 (baseline) |  |  | 1.00 (baseline) |  |  |
| **Moderate** | 1.07 | [0.75, 1.51] | 0.72 | 1.43 | [0.76, 2.67] | 0.27 |
| **Heavy** | 2.65 | [1.66, 4.22] | 0.000040 | 2.75 | [1.33, 5.71] | 0.0065 |
| **Non-binge** | 1.00 (baseline) |  |  | 1.00 (baseline) |  |  |
| **Binge** | 2.31 | [1.29, 4.14] | 0.0050 | 2.01 | [1.42, 2.85] | 0.000090 |
| **2011** |  |  |  |  |  |  |
| **Abstainer** | 2.24 | [1.27, 3.94] | 0.0051 | 3.48 | [1.02, 11.88] | 0.046 |
| **Light** | 1.00 (baseline) |  |  | 1.00 (baseline) |  |  |
| **Moderate** | 1.68 | [1.18, 2.40] | 0.0041 | 1.86 | [0.88, 3.92] | 0.11 |
| **Heavy** | 2.38 | [1.58, 3.58] | 0.000030 | 2.51 | [1.22, 5.16] | 0.012 |
| **Non-binge** | 1.00 (baseline) |  |  | 1.00 (baseline) |  |  |
| **Binge** | 1.64 | [1.17, 2.31] | 0.0044 | 1.51 | [0.86, 2.65] | 0.16 |
| **Short sleep** |  |  |  |  |  |  |
| **1975** |  |  |  |  |  |  |
| **Abstainer** | 1.34 | [0.72, 2.47] | 0.36 | 1.05 | [0.37, 2.99] | 0.92 |
| **Light** | 1.00 (baseline) |  |  | 1.00 (baseline) |  |  |
| **Moderate** | 1.12 | [0.77, 1.62] | 0.56 | 1.09 | [0.63, 1.89] | 0.76 |
| **Heavy** | 2.27 | [1.36, 3.78] | 0.0021 | 2.23 | [1.06, 4.66] | 0.034 |
| **Non-binge** | 1.00 (baseline) |  |  | 1.00 (baseline) |  |  |
| **Binge** | 1.36 | [0.98, 1.89] | 0.068 | 0.88 | [0.56, 1.38] | 0.58 |
| **1981** |  |  |  |  |  |  |
| **Abstainer** | 1.03 | [0.64, 1.66] | 0.91 | 1.37 | [0.60, 3.12] | 0.45 |
| **Light** | 1.00 (baseline) |  |  | 1.00 (baseline) |  |  |
| **Moderate** | 1.26 | [0.93, 1.71] | 0.14 | 1.11 | [0.73, 1.67] | 0.62 |
| **Heavy** | 2.09 | [1.29, 3.39] | 0.0029 | 1.69 | [0.89, 3.19] | 0.11 |
| **Non-binge** | 1.00 (baseline) |  |  | 1.00 (baseline) |  |  |
| **Binge** | 1.33 | [0.98, 1.80] | 0.064 | 1.36 | [0.88, 2.08] | 0.16 |
| **1990** |  |  |  |  |  |  |
| **Abstainer** | 0.82 | [0.45, 1.49] | 0.52 | 0.56 | [0.12, 2.58] | 0.46 |
| **Light** | 1.00 (baseline) |  |  | 1.00 (baseline) |  |  |
| **Moderate** | 1.01 | [0.70, 1.45] | 0.96 | 0.70 | [0.38, 1.31] | 0.27 |
| **Heavy** | 1.39 | [0.84, 2.31] | 0.20 | 0.82 | [0.38, 1.77] | 0.62 |
| **Non-binge** | 1.00 (baseline) |  |  | 1.00 (baseline) |  |  |
| **Binge** | 1.25 | [0.85, 1.82] | 0.26 | 1.42 | [0.82, 2.45] | 0.21 |
| **2011** |  |  |  |  |  |  |
| **Abstainer** | 1.63 | [0.92, 2.89] | 0.095 | 1.31 | [0.42, 4.07] | 0.64 |
| **Light** | 1.00 (baseline) |  |  | 1.00 (baseline) |  |  |
| **Moderate** | 1.34 | [0.97, 1.86] | 0.077 | 1.23 | [0.69, 2.18] | 0.48 |
| **Heavy** | 1.64 | [1.08, 2.49] | 0.019 | 1.34 | [0.68, 2.63] | 0.40 |
| **Non-binge** | 1.00 (baseline) |  |  | 1.00 (baseline) |  |  |
| **Binge** | 1.27 | [0.94, 1.72] | 0.12 | 1.25 | [0.72, 2.20] | 0.43 |

Model 1 includes sex and age as covariates, whereas Model 2 was adjusted for sex, age, BMI, smoking and life satisfaction. Statistically significant (P<0.05) associations are highlighted in red.

*Total N includes all dizygotic twin participants who have taken at least one of the four surveys and answered some, but not necessarily all, of the questions of that survey.
